# Metagenomic next-generation sequencing of cerebrospinal fluid for diagnosis of central nervous system infections: 7-year performance of a clinically validated test

**DOI:** 10.1101/2024.03.14.24304139

**Authors:** Patrick Benoit, Noah Brazer, Emily Kelly, Venice Servellita, Miriam Oseguera, Jenny Nguyen, Jack Tang, Mikael de Lorenzi-Tognon, Charles Omura, Jessica Streithorst, Melissa Hillberg, Danielle Ingebrigtsen, Kelsey Zorn, Michael Wilson, Tim Blicharz, Amy P. Wong, Brian O’Donovan, Brad Murray, Steve Miller, Charles Y. Chiu

## Abstract

Studies have shown that metagenomic next-generation sequencing (mNGS) testing of cerebrospinal fluid (CSF) in central nervous system (CNS) infections can improve diagnostic yields and provide actionable information. We analyzed the results of all CSF mNGS tests (n=4,828) performed at the University of California, San Francisco (UCSF) clinical microbiology laboratory from June 2016 to April 2023. We also assessed clinical metadata from a subset of samples that corresponded to a cohort of UCSF patients (n=1,164) who received CSF mNGS testing, and retrospectively evaluated performance compared to conventional microbiologic testing and adjudicated clinical diagnosis. Overall, 14.4% of CSF mNGS tests were positive for any microorganism. DNA viruses (7% of all samples) were detected most often, followed by RNA viruses (4.3%), bacteria (2.7%), fungi (1.4%), and parasites (0.5%). Using a composite gold standard obtained from clinical adjudication and all microbiological test results, sensitivity, specificity, and accuracy of CSF mNGS in the UCSF cohort who had clinically diagnosed infections were 56.5%, 98.8%, and 90.5%, respectively. The sensitivity of CSF mNGS testing (56.5%) was statistically higher than that from all direct detection testing from CSF (44.8%, p = 0.004), direct detection testing from samples other than CSF (15.2%, p<0.001), and indirect serologic testing (34%, p<0.001). When only considering diagnoses made by direct detection of pathogens on CSF, sensitivity of mNGS was 80.7%. These results justify the incorporation of CSF mNGS testing as part of the routine diagnostic workup in hospitalized patients presenting with potential CNS infection.

## Introduction

Central nervous system (CNS) infections associated with meningitis, encephalitis, and/or myelitis can cause severe and often life-threatening illness^1,2^. Timely diagnosis and rapid initiation of appropriate therapy are paramount to improve clinical outcomes in these infections, and delays have been associated with increased morbidity and mortality in cases of bacterial meningitis and herpes simplex virus encephalitis^3,4^. A complete diagnostic workup for CNS infection includes a combination of culture-based, serologic, and nucleic acid amplification tests (NAAT). However, despite extensive testing, it is estimated that the cause of meningoencephalitis remains unknown in around 50% of cases, which hinders the initiation of appropriate and targeted therapy^5,6^.

In recent years, clinical metagenomic next-generation sequencing (mNGS) has emerged as a comprehensive approach for infectious disease diagnosis, enabling simultaneous detection of a wide range of microorganisms, including bacteria, viruses, fungi, and parasites, without the need to target a specific pathogen, as is required in routine microbiological testing^7,8^. This agnostic diagnostic method can be particularly useful in CNS infections when the differential diagnosis is often very broad, with overlapping clinical manifestations of infectious and non-infectious etiologies, and when there is limited availability of CNS samples such as cerebrospinal fluid (CSF) and brain biopsy. Studies have previously shown that CSF mNGS testing in CNS infections can improve diagnostic yields and provide clinically actionable information^6^.

The University of California in San Francisco (UCSF) CSF mNGS test (“UCSF mNGS test”) was developed in 2016; the test was analytically and clinically validated as a laboratory-developed test and is conducted in a CLIA-certified laboratory^9^. The UCSF mNGS test sequences both DNA and RNA for a comprehensive evaluation of infectious pathogens, and has been integrated into UCSF’s testing algorithm for cases of suspected CNS infection. However, the performance of the test longitudinally and in real-life clinical settings has yet to be reported. Here, we review the results from 4,828 samples tested by CSF mNGS from 2016 to 2023 to evaluate (i) quality control (QC) metrics, (ii) turnaround times, and (iii) detection yields for microorganisms. We also assess a subset of 1,164 samples from UCSF patients (“UCSF cohort”) by conducting a detailed chart review to retrospectively evaluate test performance compared to conventional testing and adjudicated clinical diagnosis.

## Results

### UCSF mNGS test cohort description

Overall, 4,828 CSF mNGS tests were performed by the UCSF clinical microbiology laboratory from June 2016 to April 2023, including testing of 1,130 UCSF patients **(Table 1)**. The number of tests performed increased by nearly 10-fold from 116 in 2016 to 1,067 in 2022 **(Figure 1)**. 55.7% of patients were male; the mean age was 41.5 years old with 24.2% of the cohort younger than 18 years old. Most CSF mNGS tests were performed for US patients representing 46 different states (n=4,075), with 31 tests for patients from other countries (Canada was the most frequent). 722 samples were sent from national or regional reference laboratories **(Figure 1)**. Median turnaround times for UCSF and non-UCSF patients were 8.2 and 11.4 days (p < 0.0001), respectively, from sample collection to result, and 3.6 and 3.8 days, respectively, from start of sample processing in the laboratory to result (p < 0.0001). The longer turnaround times from sample collection to test results for non-UCSF patients may be explained by the delays between the lumbar puncture and decision to send the sample for mNGS testing, delays from shipping of samples, and delays from batching of samples prior to testing.

**Figure 1.**
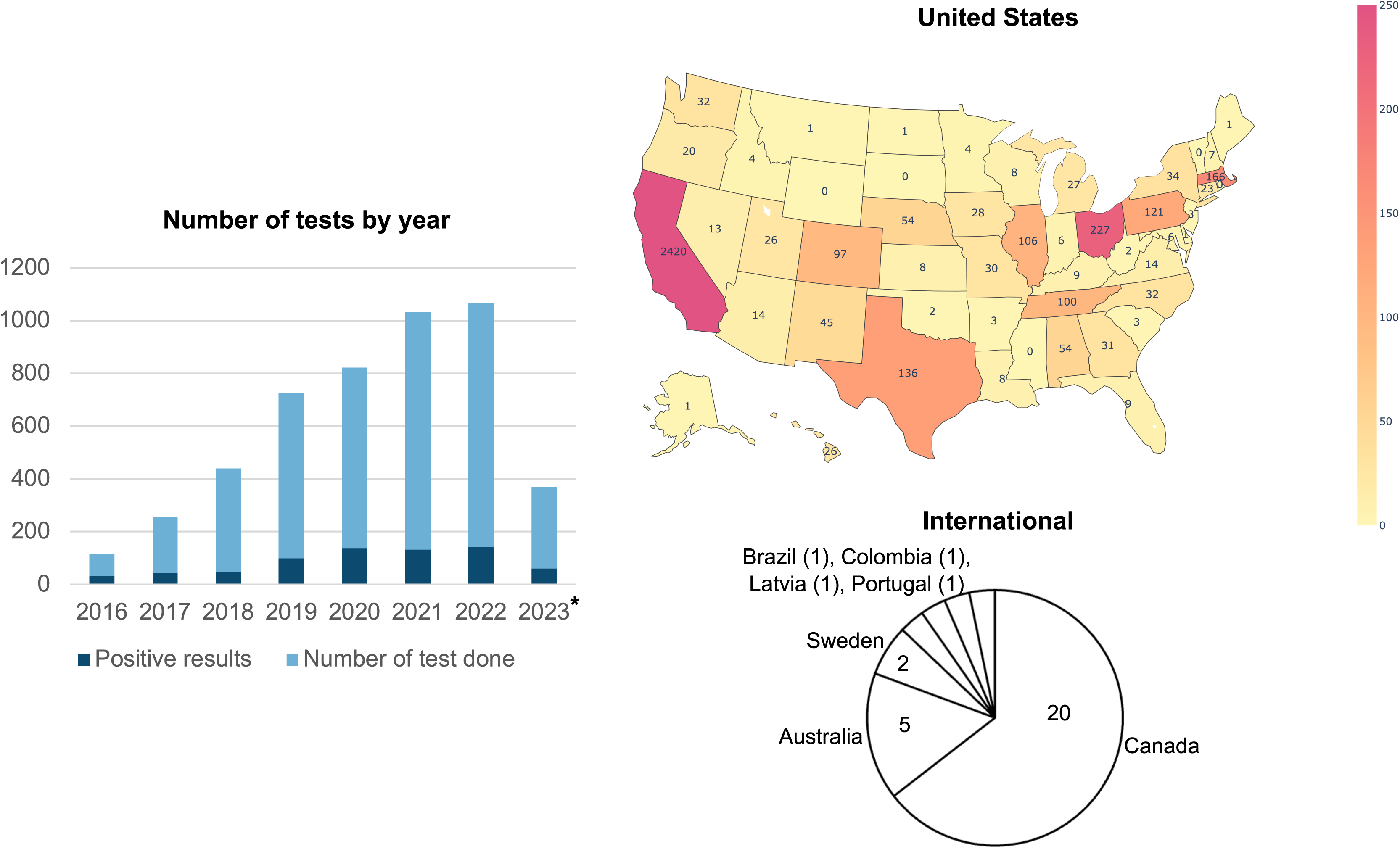
Distribution of tests ordered by year and by location. Distribution of tests ordered by State, internationally, and from reference laboratories. A total of 4075 tests were from the United States, California being the most frequent State of origin (n = 2420 samples). Reference laboratories such as ARUP, Labcorp, and Mayo Clinic Laboratories (n=722) receive tests from multiple States, so individual samples couldn’t be retraced and, therefore, excluded from the figure. 14.8 % (n = 715) of samples were sent from hospitals dedicated to the care of pediatric patients. * Data are up to April 2023.

**Table 1.**
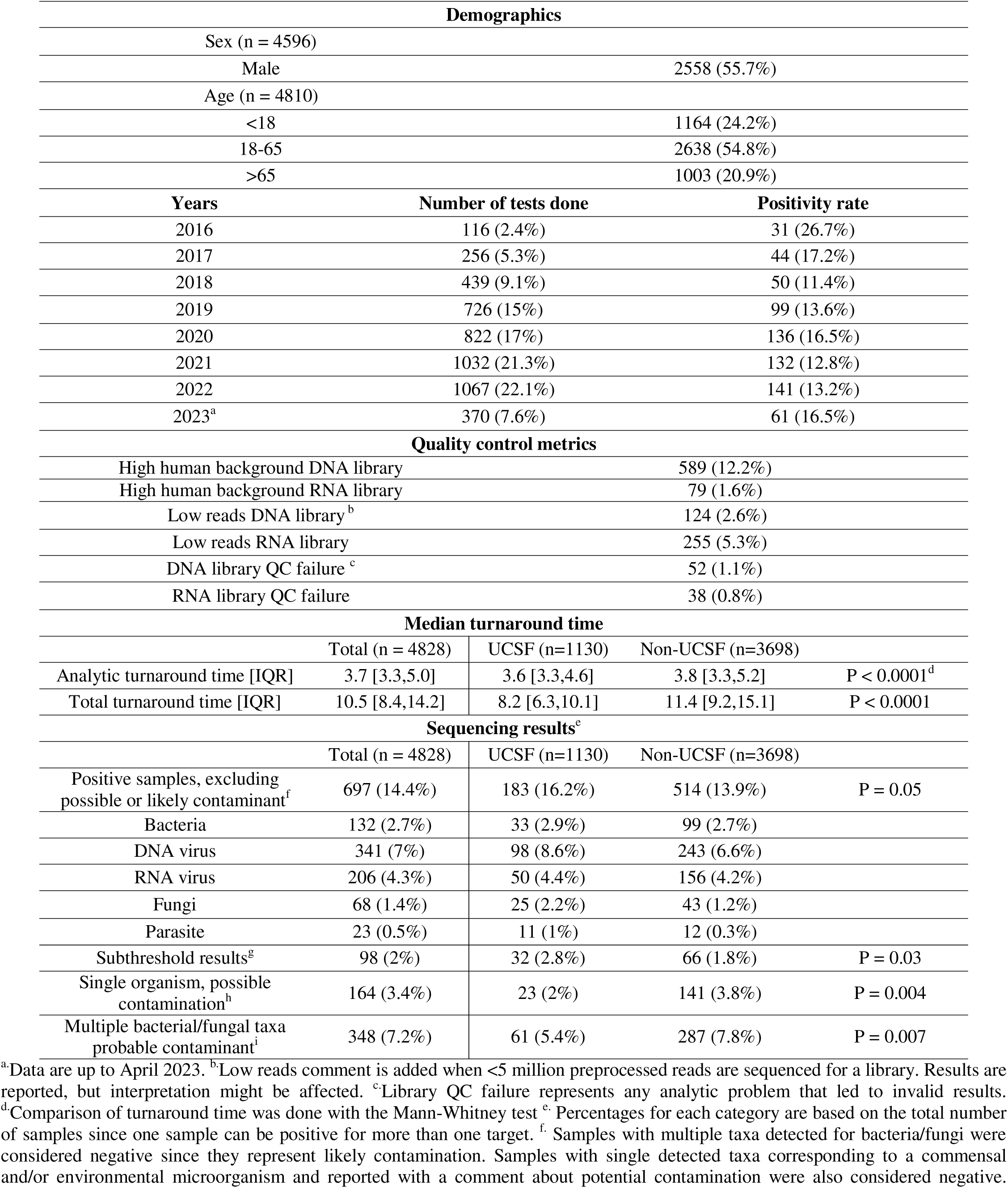
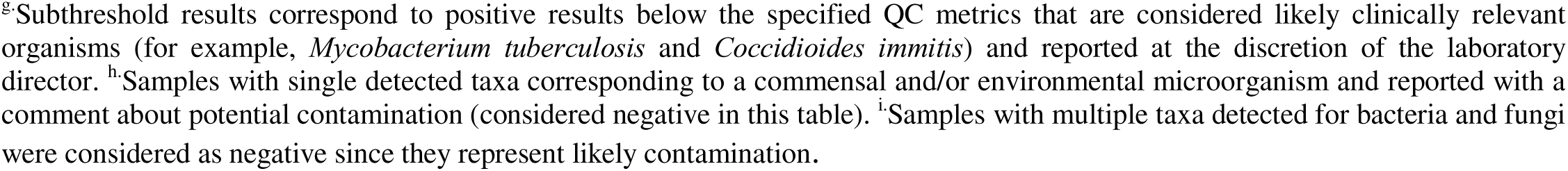
UCSF mNGS test characteristics for 4828 samples performed from 2016 to 2023.

| Demographics |  |  |  |  |
| --- | --- | --- | --- | --- |
| Sex (n = 4596) |  |  |  |  |
| Male |  |  | 2558 (55.7%) |  |
| Age (n = 4810) |  |  |  |  |
| <18 |  |  | 1164 (24.2%) |  |
| 18-65 |  |  | 2638 (54.8%) |  |
| >65 |  |  | 1003 (20.9%) |  |
| Years | Number of tests done |  | Positivity rate |  |
| 2016 | 116 (2.4%) |  | 31 (26.7%) |  |
| 2017 | 256 (5.3%) |  | 44 (17.2%) |  |
| 2018 | 439 (9.1%) |  | 50 (11.4%) |  |
| 2019 | 726 (15%) |  | 99 (13.6%) |  |
| 2020 | 822 (17%) |  | 136 (16.5%) |  |
| 2021 | 1032 (21.3%) |  | 132 (12.8%) |  |
| 2022 | 1067 (22.1%) |  | 141 (13.2%) |  |
| 2023<sup>a</sup> | 370 (7.6%) |  | 61 (16.5%) |  |
| Quality control metrics |  |  |  |  |
| High human background DNA library |  |  | 589 (12.2%) |  |
| High human background RNA library |  |  | 79 (1.6%) |  |
| Low reads DNA library <sup>b</sup> |  |  | 124 (2.6%) |  |
| Low reads RNA library |  |  | 255 (5.3%) |  |
| DNA library QC failure <sup>c</sup> |  |  | 52 (1.1%) |  |
| RNA library QC failure |  |  | 38 (0.8%) |  |
| Median turnaround time |  |  |  |  |
|  | Total (n = 4828) | UCSF (n=1130) | Non-UCSF (n=3698) |  |
| Analytic turnaround time [IQR] | 3.7 [3.3,5.0] | 3.6 [3.3,4.6] | 3.8 [3.3,5.2] | P < 0.0001<sup>d</sup> |
| Total turnaround time [IQR] | 10.5 [8.4,14.2] | 8.2 [6.3,10.1] | 11.4 [9.2,15.1] | P < 0.0001 |
| Sequencing results<sup>e</sup> |  |  |  |  |
|  | Total (n = 4828) | UCSF (n=1130) | Non-UCSF (n=3698) |  |
| Positive samples, excluding possible or likely contaminant<sup>f</sup> | 697 (14.4%) | 183 (16.2%) | 514 (13.9%) | P = 0.05 |
| Bacteria | 132 (2.7%) | 33 (2.9%) | 99 (2.7%) |  |
| DNA virus | 341 (7%) | 98 (8.6%) | 243 (6.6%) |  |
| RNA virus | 206 (4.3%) | 50 (4.4%) | 156 (4.2%) |  |
| Fungi | 68 (1.4%) | 25 (2.2%) | 43 (1.2%) |  |
| Parasite | 23 (0.5%) | 11 (1%) | 12 (0.3%) |  |
| Subthreshold results<sup>g</sup> | 98 (2%) | 32 (2.8%) | 66 (1.8%) | P = 0.03 |
| Single organism, possible contamination<sup>h</sup> | 164 (3.4%) | 23 (2%) | 141 (3.8%) | P = 0.004 |
| Multiple bacterial/fungal taxa probable contaminant<sup>i</sup> | 348 (7.2%) | 61 (5.4%) | 287 (7.8%) | P = 0.007 |

<sup>a</sup>Data are up to April 2023. <sup>b</sup>Low reads comment is added when <5 million preprocessed reads are sequenced for a library. Results are reported, but interpretation might be affected. <sup>c</sup>Library QC failure represents any analytic problem that led to invalid results. <sup>d</sup>Comparison of turnaround time was done with the Mann-Whitney test <sup>e</sup> Percentages for each category are based on the total number of samples since one sample can be positive for more than one target. <sup>f</sup> Samples with multiple taxa detected for bacteria/fungi were considered negative since they represent likely contamination. Samples with single detected taxa corresponding to a commensal and/or environmental microorganism and reported with a comment about potential contamination were also considered negative.
<sup>g</sup>Subthreshold results correspond to positive results below the specified QC metrics that are considered likely clinically relevant organisms (for example, *Mycobacterium tuberculosis* and *Coccidioides immitis*) and reported at the discretion of the laboratory director. <sup>h</sup>Samples with single detected taxa corresponding to a commensal and/or environmental microorganism and reported with a comment about potential contamination (considered negative in this table). <sup>i</sup>Samples with multiple taxa detected for bacteria and fungi were considered as negative since they represent likely contamination.

We also evaluated QC metrics and microorganism detection yields associated with CSF mNGS testing. High host background associated with presumptive reduced assay sensitivity^10^ was more frequently seen in DNA libraries (12.2%) than in RNA libraries (1.6%). This result illustrates the interference of host DNA background and the higher efficiency of DNase treatment (RNA libraries) compared to antibody-based methylated DNA removal (DNA libraries) in reducing host DNA background. On the other hand, RNA libraries were more difficult to amplify due to low read counts which were more frequent for RNA libraries (5.3%) than for DNA libraries (2.6%). QC failure due to inadvertent errors in sample processing that required additional repeat testing was seen rarely (<~1%).

Out of 4,828 samples, 697 samples (14.4%) were positive for any microorganism, excluding possible or likely contaminants **(Table 1, Supplementary Dataset 1).** The annual positivity rate varied from 11.4% to 17,4%. Notably, for 2016, the positivity rate was 27%, which corresponds to the year the prospective Precision Diagnosis of Acute Infectious Diseases (PDAID) study was conducted using enrollment criteria to select patients for mNGS testing^6^. DNA viruses were most frequently identified (7% of all samples), followed by RNA viruses (4.3%), bacteria (2.7%), fungi (1.4%) and parasites (0.5%). Multiple bacterial and/or fungal taxa were detected in 7.2% of samples (348/4828) and single organisms corresponding to a commensal and/or environmental species in 3.4% of samples (164/4828). These results categories were reported more frequently for patients outside of UCSF (7.8% vs. 5.4 %, p=0.007 and 3.8% vs. 2%, p=0.004, respectively) and can be attributed to differences in CSF collection or transportation methods. These results were classified as negative since they are clinically considered likely to represent sample contamination rather than detection of etiology, as indicated in the clinical report^6^. The positivity rate in UCSF (16.2%) was higher than in non-UCSF (13.9%) patients (p=0.05). This difference can be explained in part by the higher rate of results for UCSF patients that were subthreshold (2.9% vs. 1.8%, p=0.03) (the number of reads was lower than threshold values specified in the QC metrics), and were classified as positives, based on review of patient charts, clinical presentation, and discussion with the UCSF clinical team. Overall, 2% (98/4828) positive results were reported as subthreshold. 92.3% (12/13) of *Mycobacterium tuberculosis* and 93.4 % (15/16) *Coccidioides spp*. results were reported as subthreshold. Other overrepresented organisms in this category were *Balamuthia mandrillaris*, *Histoplasma capsulatum*, West Nile virus, and Powassan virus, with 66% (2/3), 50% (2/4), 28.6% (8/28), 21.4% (3/14) of their results being reported as subthreshold, respectively **(Supplementary dataset 1)**.

The pathogens detected with the UCSF mNGS test represented a large diversity of organisms (**Figure 2A**). A total of 59 unique bacterial species were detected among the 132 positive results for bacteria **(Figure 2B).** The assay detected 86 non-fastidious bacterial pathogens with 35 unique species, corresponding to bacteria that traditional culture methods can detect. 46 uncommon and/or difficult-to-diagnose pathogens from 24 unique species were detected such as *Mycobacterium tuberculosis*, *Nocardia farcinia*, *Leptospira borgpetersenii*, *Borrelia miyamotoi*, and *Tropheryma whipplei,* corresponding to rare but definitive CNS pathogens, that are fastidious or not often detected by culture (**Supplementary table 1**). The most common RNA viruses detected were HIV (92), vector-borne viruses (57), and enteroviruses (16) **(Figure 2C)**. Uncommon arboviruses **(Figure 2D)** such as St-Louis encephalitis virus, La Crosse virus, Cache Valley virus, and even a novel pathogen not previously described in humans, Potosi virus ^11^, were detected. Various serotypes of enteroviruses could also be detected, including A71 and D68, associated with acute flaccid myelitis (**Figure 2E**). Other interesting pathogens detected include *Coccidioides immitis and posadasi* (16), *Cryptococcus neoformans and gattii* (13), *Histoplasma capsulatum* (4), and *Fusarium spp*. (3) for fungi **(Figure 2G)**, and *Toxoplasma gondii* (10), *Balamuthia mandrillaris* (3), *Angiostrongylus cantonensis* (2), and *Naegleria fowleri* (1) for parasites **(Figure 2H)**. Some results were also related to the initiation of outbreak investigations. For example, a vaccine strain of Yellow fever virus in CSF from a transplant recipient with encephalitis was reported - this case prompted an investigation by the US Centers for Disease Control of a nationwide transplant-associated outbreak^12^. Also, the UCSF mNGS test uncovered an outbreak of fungal meningitis with *Fusarium solani* in the US associated with surgical procedures performed under epidural anesthesia in Matamoros, Mexico^13^.

**Figure 2.**
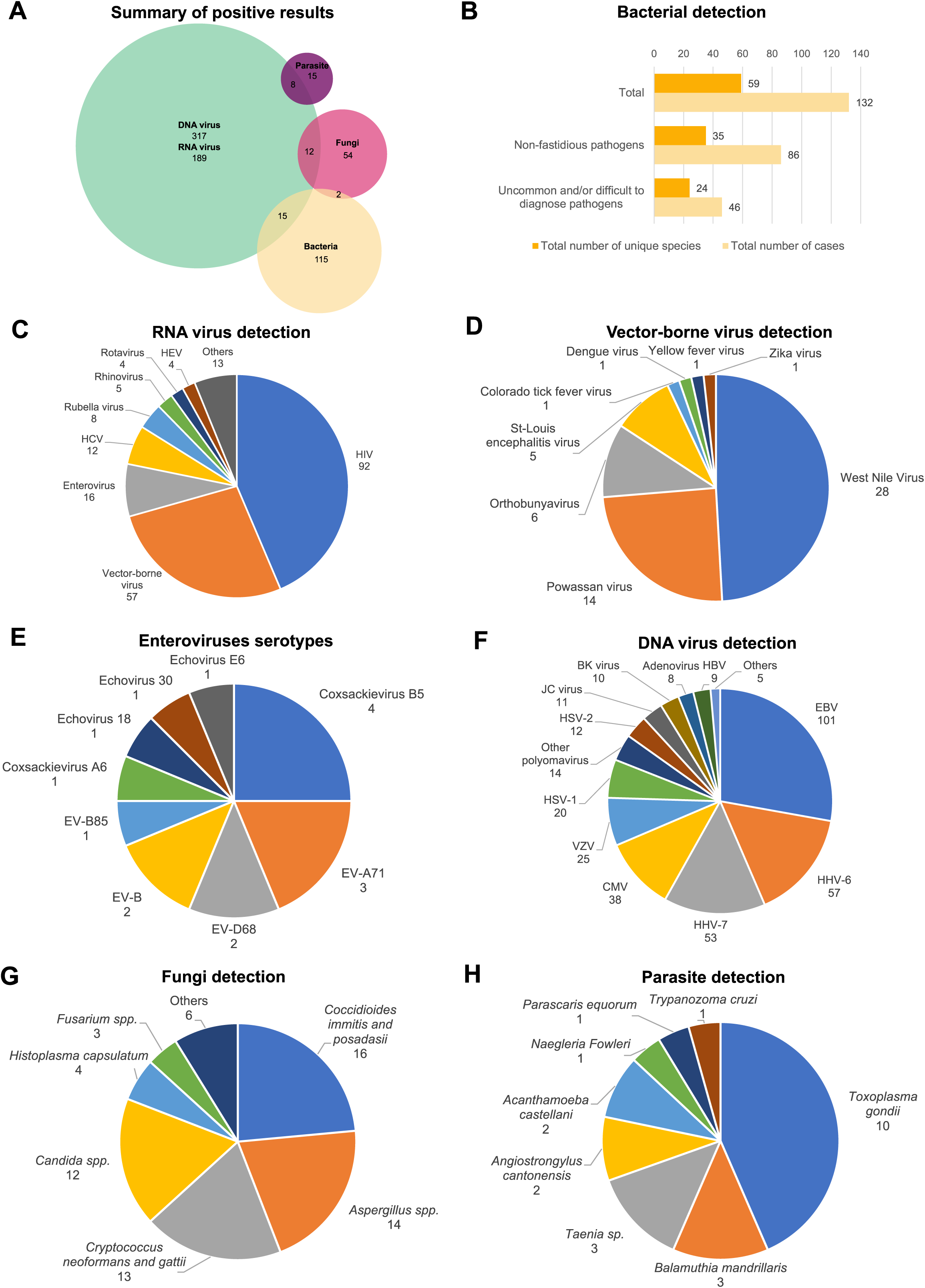
Summary of positive results by mNGS (n=4828). **(A)** Summary of positive UCSF mNGS test results by microorganism type. One test can be positive for more than one target. **(B)** A total of 59 unique species with a total of 132 cases were detected by the assay. 88 pathogens that can be detected by traditional culture methods corresponding to 35 unique species were detected. 46 uncommon and/or difficult-to-diagnose bacteria, corresponding to rare but definitive CNS pathogens, that are fastidious or not detected by culture. The list of bacterial pathogens detected in available in **supplementary table 1**. **(C)** The most common RNA viruses detected were HIV (92), vector-borne viruses (57), and enteroviruses (16). Other RNA viruses detected include Lymphocytic Choriomeningitis virus (3), Astrovirus (2), Calicivirus (2), Coronavirus 229E (2), SARS-CoV-2 (1), HTLV-2 (1), Human parechovirus (1), Measles virus (1). **(D)** Vector-borne viruses, a diverse and clinically important group, are typically challenging to diagnose by traditional methods because they require specific NAAT or serologies for each virus, whereas mNGS can detect a broad range of pathogens in a single assay. *Orthobunyaviruses* detected included Cache Valley virus (3), Jamestown Canyon virus (1) La Crosse virus (1), Potosi virus (1). Of interest, the Potosi virus is a novel species in this genus, not reported before as causing diseases in humans ^11^. **(E)** One of the strengths of the assay is its ability to discriminate between species and serotypes, unlike the typical testing method of NAAT. For instance, the assay detected various subtypes of enteroviruses and five cases of A71 and D68 associated with acute flaccid myelitis. **(F)** Other DNA viruses detected included Human parvovirus 4 and B-19 (4) and HHV-8 (1). **(G)** Other fungi detected included *Alternaria spp.* (2), *Mucorales spp.* (2)., *Epicoccum spp.* (1), and *Sporothrix schenkii* (1). **(H)** The assay detected challenging parasitic infections to diagnose, such as *Toxoplasma gondii* (10), *Balamuthia mandrillaris* (3), *Angiostrongylus cantonensis* (2), and *Naegleria fowleri* (1). HCV: Hepatitis C virus, HEV: Hepatitis E virus, HIV: Human immunodeficiency virus, EV: Enterovirus, EBV: Epstein-Barr virus, CMV: Cytomegalovirus, HHV-6: Human herpes virus 6, HHV-7: Human herpes virus 7, VZV: Varicella-zoster virus, HSV: Herpes simplex virus, NAAT: Nucleic acid amplification test.

### UCSF patient cohort

Results from 1,164 CSF samples tested by mNGS from June 2016 to June 2023 were analyzed along with a retrospective chart review of the corresponding 1052 patients (some patients had multiple CSF mNGS tests performed). The UCSF cohort consisted of 223 (19.2%) cases of microbiologically confirmed CNS infection, 431 (37%) cases of autoimmune disease or another presumably non-infectious condition, 1 case (0.1%) of prion disease, and 509 (43.7%) cases of meningitis/encephalitis with unknown etiology despite all diagnostic testing including mNGS testing and longitudinal follow-up (**Table 2**). Among all UCSF patients who received the UCSF mNGS test, 425 (36.5%) were immunocompromised, 1021 (87.7%) were hospitalized, 450 (38.7%) had an intensive care unit (ICU) stay with a with a mean length of stay of 21.6 days, and 119 (10.2%) died within 60 days of admission. The average number of diagnostic tests performed was 6 from CSF and 20.2 from all samples (including CSF), consisting of 28.2% (average of 5.7) culture, 24.8% (5) NAAT, 13.4% (2.7) antigen, and 33.7% (6.8) serology-based assays.

**Table 2.**
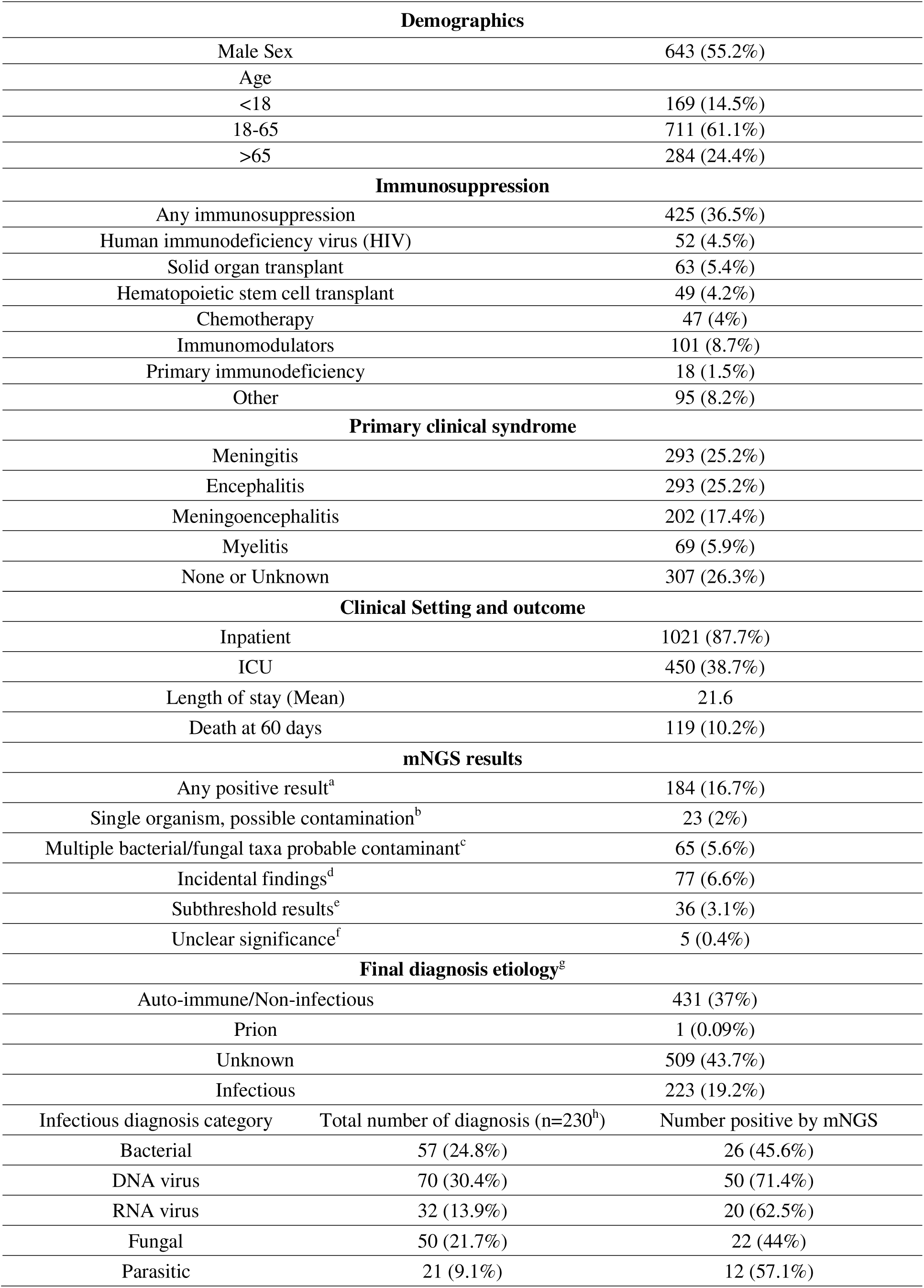
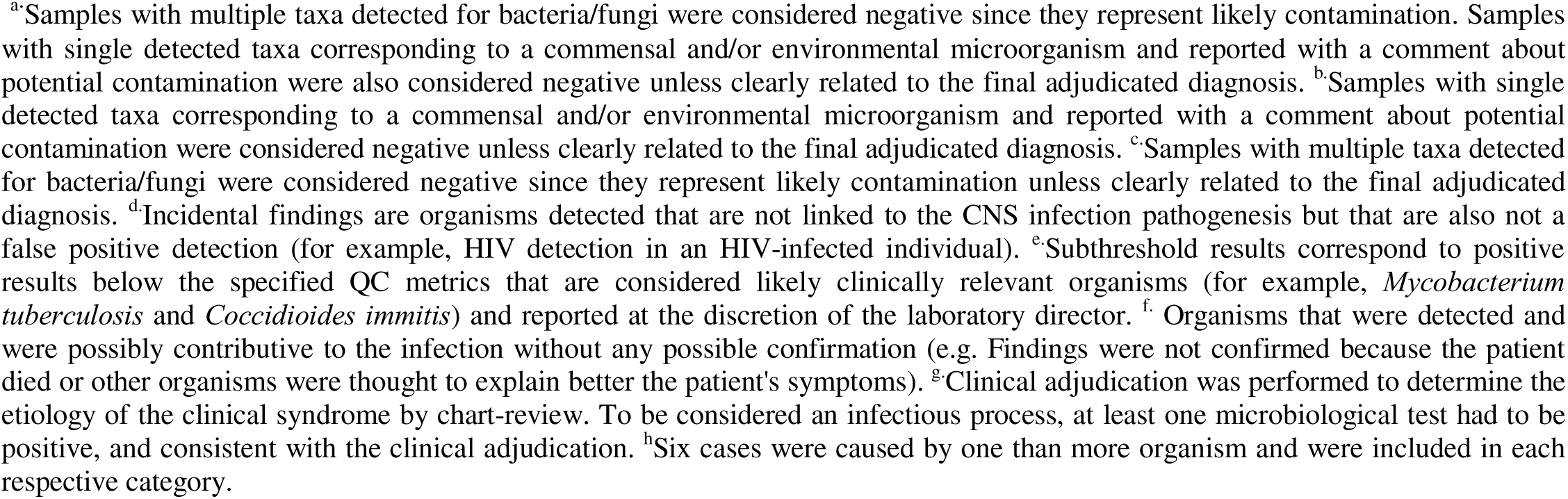
Subset analysis from the UCSF cohort (n = 1164).

| Demographics |  |  |
| --- | --- | --- |
| Male Sex |  | 643 (55.2%) |
| Age |  |  |
| <18 |  | 169 (14.5%) |
| 18-65 |  | 711 (61.1%) |
| >65 |  | 284 (24.4%) |
| Immunosuppression |  |  |
| Any immunosuppression |  | 425 (36.5%) |
| Human immunodeficiency virus (HIV) |  | 52 (4.5%) |
| Solid organ transplant |  | 63 (5.4%) |
| Hematopoietic stem cell transplant |  | 49 (4.2%) |
| Chemotherapy |  | 47 (4%) |
| Immunomodulators |  | 101 (8.7%) |
| Primary immunodeficiency |  | 18 (1.5%) |
| Other |  | 95 (8.2%) |
| Primary clinical syndrome |  |  |
| Meningitis |  | 293 (25.2%) |
| Encephalitis |  | 293 (25.2%) |
| Meningoencephalitis |  | 202 (17.4%) |
| Myelitis |  | 69 (5.9%) |
| None or Unknown |  | 307 (26.3%) |
| Clinical Setting and outcome |  |  |
| Inpatient |  | 1021 (87.7%) |
| ICU |  | 450 (38.7%) |
| Length of stay (Mean) |  | 21.6 |
| Death at 60 days |  | 119 (10.2%) |
| mNGS results |  |  |
| Any positive result<sup>a</sup> |  | 184 (16.7%) |
| Single organism, possible contamination<sup>b</sup> |  | 23 (2%) |
| Multiple bacterial/fungal taxa probable contaminant<sup>c</sup> |  | 65 (5.6%) |
| Incidental findings<sup>d</sup> |  | 77 (6.6%) |
| Subthreshold results<sup>e</sup> |  | 36 (3.1%) |
| Unclear significance<sup>f</sup> |  | 5 (0.4%) |
| Final diagnosis etiology<sup>g</sup> |  |  |
| Auto-immune/Non-infectious |  | 431 (37%) |
| Prion |  | 1 (0.09%) |
| Unknown |  | 509 (43.7%) |
| Infectious |  | 223 (19.2%) |
| Infectious diagnosis category | Total number of diagnosis (n=230<sup>h</sup>) | Number positive by mNGS |
| Bacterial | 57 (24.8%) | 26 (45.6%) |
| DNA virus | 70 (30.4%) | 50 (71.4%) |
| RNA virus | 32 (13.9%) | 20 (62.5%) |
| Fungal | 50 (21.7%) | 22 (44%) |
| Parasitic | 21 (9.1%) | 12 (57.1%) |

For each case, we established a composite diagnosis that incorporated clinical adjudication with the results of all microbiologic testing (**Supplementary dataset 2**). There were 184 (15.8%) cases of positive detection of a microorganism by CSF mNGS, of which 36 (3.1%) were considered as subthreshold positive results **(Table 2)**. 77 samples (6.6%) included incidental findings (for example, HIV detection in an HIV-infected patient with an acute neurologic illness not explained by HIV) and were excluded from further analysis. 5 samples (0.4%) included results of unclear significance and were also excluded (1 sample with HSV-1, 2 with CMV, 1 with LCMV, and 1 with *Candida albicans*). Among results with a comment about possible contamination and multiple bacterial/fungal taxa, only 2/22 (9.1%) and 1/65 (1.5%) were considered positive upon clinical review, respectively, as they were clearly related to the final adjudicated diagnosis **(Supplementary table 5 and 6)**. Among the included mNGS positive results, 130 were true positive, where a target relevant to the adjudicated diagnosis was detected, and 11 were false positive, detecting a target not relevant to the final diagnosis **(Table 3).** For the false positive results, none of the final diagnoses were due to an infectious etiology, and none of the patients received treatment for the organism detected. Most of the false positive results were bacteria (75%) **(Figure 4, Supplementary table 4)**. 86% (31/36) of subthreshold results were considered true positive. Among the mNGS negative results, 930 were true negative and 100 were false negative. Overall, CSF mNGS testing exhibited a sensitivity of 56.5%, specificity of 98.8%, accuracy of 90.5%, positive predictive value (PPV) of 92.2% and negative predictive value (NPV) of 90.3% for CNS infections (**Table 3**). The sensitivity of CSF mNGS (56.5%) was statistically higher than all other diagnostic modalities, including direct detection tests from CSF (44.8%, p=0.004), direct detection tests from samples other than CSF (16.2%, p<0.001), and indirect serologic detection from blood (or blood components including serum and plasma) or CSF (30.3%, p<0.001) **(Table 3).** A proportional Venn diagram revealed that 46 infectious diagnoses were made exclusively using the UCSF mNGS test (20% of all diagnosis), with the overlap between mNGS and other testing modalities ranging from 6 to 63 diagnoses **(Figure 3).** The UCSF mNGS test was the most sensitive diagnostic modality for RNA viruses, DNA viruses, bacteria, and parasites, but not for fungi, which was detected more often by culture and antigen detection methods. When only considering diagnosis made by direct detection of pathogen on CSF, positive percentage agreement (PPA) of mNGS was 80.7%.

**Figure 3.**
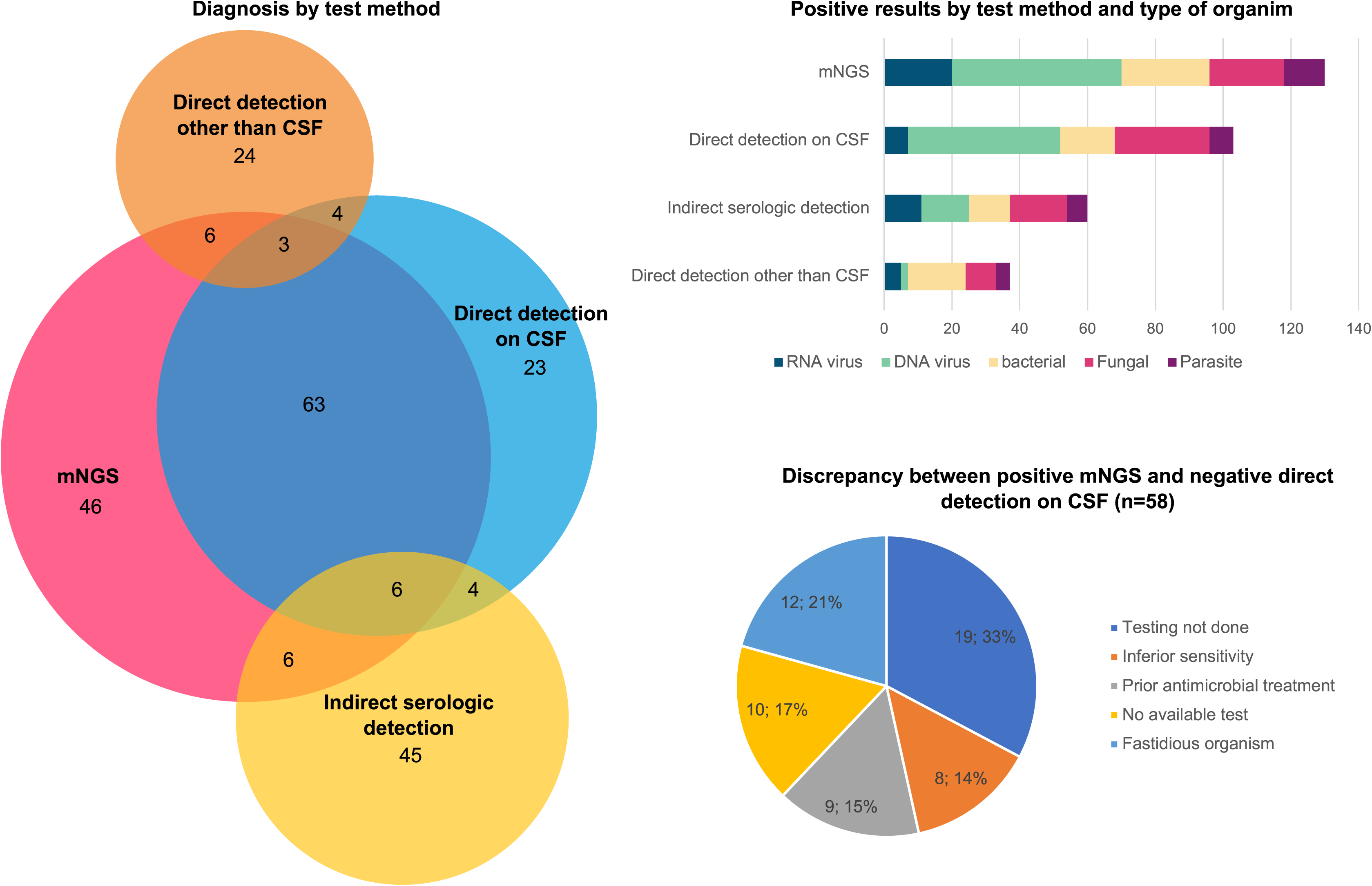
Comparison of each diagnostic modality in obtaining a final diagnosis and analysis of positive mNGS results. **(A)** The proportional Venn diagram represents the overlap between diagnostic modalities, with CSF mNGS and direct detection on CSF having the biggest overlap due to the similarity in the analytic principle (detection of pathogen). A substantial number of diagnoses were made only by indirect serologic methods (44), and direct detection on samples other than CSF (24). The total number of diagnoses is 230 (Six cases were attributed to more than one causative organism and were considered individually for this analysis). Diagnoses made by mNGS that were negative by direct detection methods (n=58) included common (Cytomegalovirus (CMV), Coxsackieviruses, *Streptococcus pneumoniae*) and rare pathogens (Lymphocytic Choriomeningitis virus, *Ureaplasma parvum*, *Balamuthia mandrillaris*). When the diagnosis was made by both mNGS and direct detection methods on CSF, 16/72 (22%) were made faster by mNGS, implying that most of these diagnoses were, in fact, made by mNGS and confirmed by a secondary analysis. These results are available in **supplementary table 2**. **(B)** CSF mNGS detected the most cases of RNA viruses, DNA viruses, bacteria, and parasites, but not for fungi, which was detected more often by culture and antigen detection methods. **(C)** The most common discrepant factor to explain an mNGS positive result and a negative direct detection method on CSF was failure to specifically test for the pathogen when a test was readily available (33%), inability to culture a fastidious micro-organism (21%), failure to specifically test for the pathogen when there is no readily available test (17%), prior antimicrobial therapy with negative culture at time of CSF collection (15%), and inferior sensitivity of conventional tests compared to mNGS (14%).

**Figure 4.**
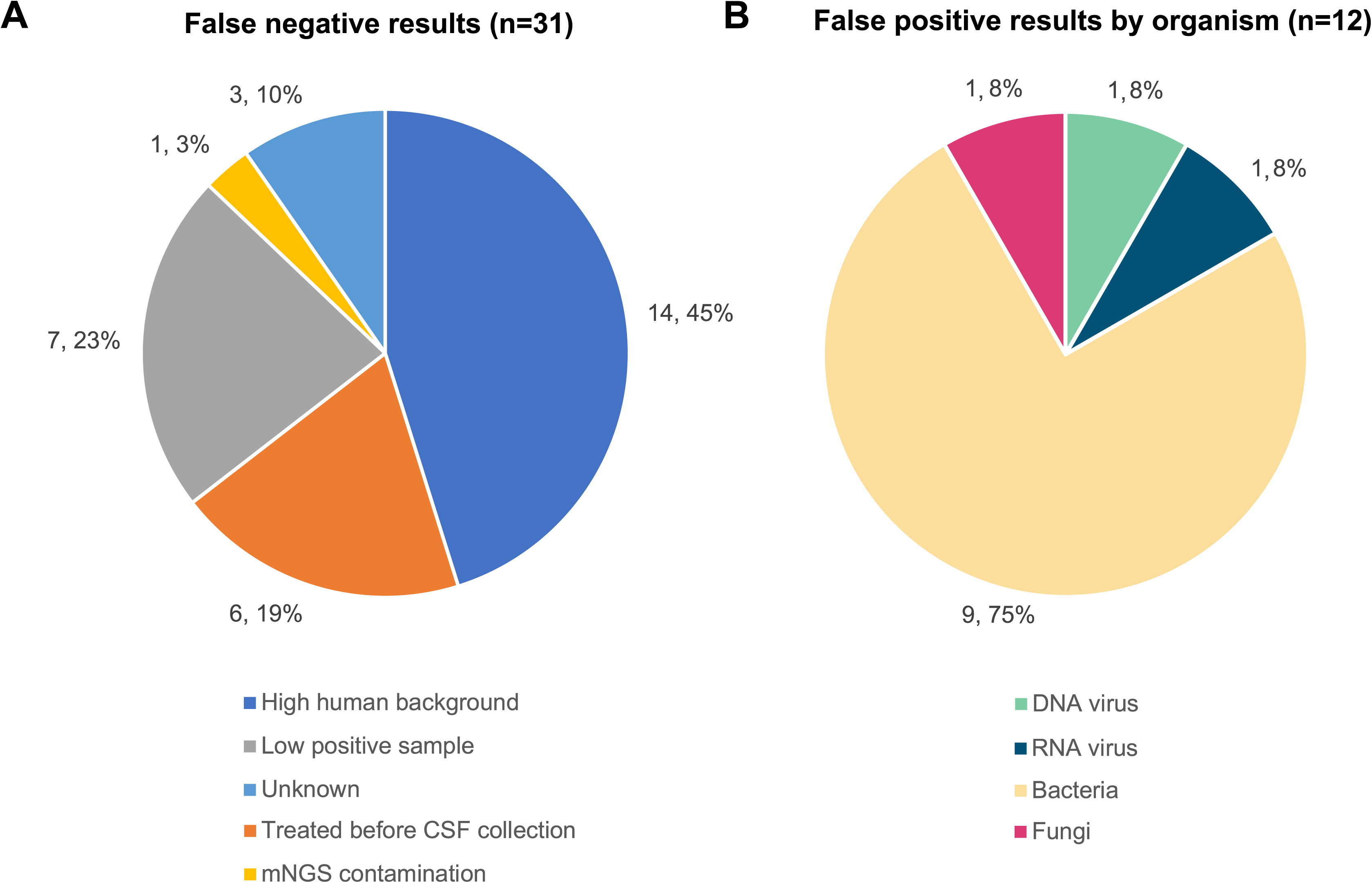
Analysis of false negative and false positive mNGS results. **(A)** 31 samples were positive by direct detection methods on CSF and negative by mNGS. 64% of the false negative results were attributed to either high DNA host background (45%), a known limitation of mNGS approaches^7,10^, or persistent antigen positivity from fungal infection after onset of treatment (19%). The list of mNGS false negative results is available in **supplementary table 3**. **(B)** False positive results were infrequent (n=11), with a majority of bacteria detected (75%). None of these mNGS results were acted upon, and all pathogens detected by mNGS were not compatible with the clinical presentation of the patient. The list of false positive results is available in **supplementary table 4**.

**Table 3.**
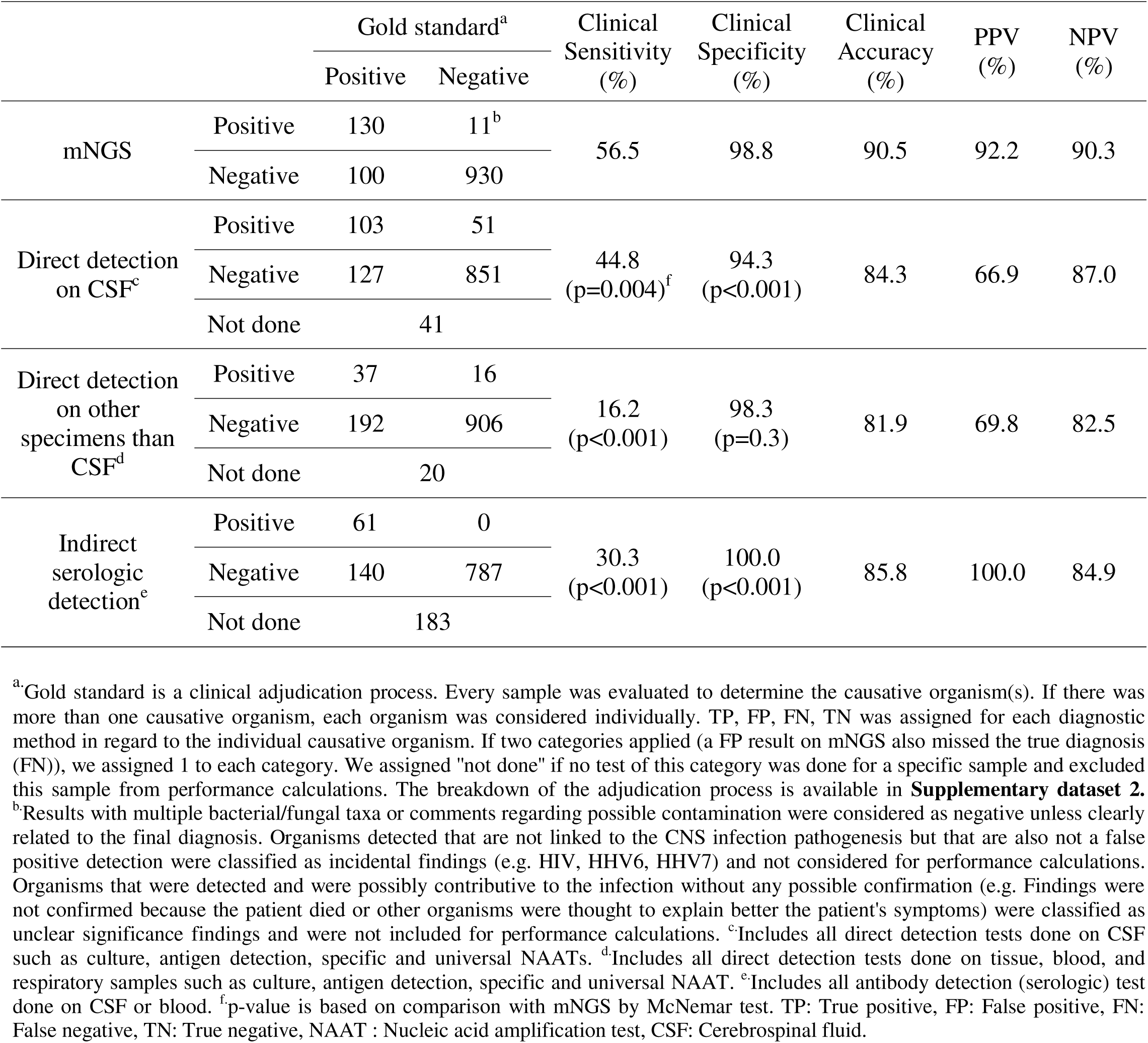
Clinical performance of diagnostic methods for diagnosis of CNS infections.

As mNGS is most comparable to combined direct detection tests from CSF in the analytic principle, we analyzed discrepant results between the two diagnostic modalities. 58 infectious diagnoses were positive by CSF mNGS and not detected by another direct detection modality (**Supplementary table 2**). 33% (19/58) were not detected because no test was ordered that specifically targeted the pathogen despite the availability of such test, 17% (10/58) were not detected because a test to detect the pathogen was not readily available (e.g., Lymphocytic Choriomeningitis Virus), 14% (8/58) were not detected because the test had lower sensitivity compared to mNGS, 21% (12/58) were not detected because of fastidious microorganisms like mycobacteria and fungi that were not detectable by culture, and 15% (9/58) were not detected because antimicrobial treatment before CSF collection led to falsely negative culture **(Figure 3)**. Among the 72 cases for which both the UCSF mNGS test and direct detection testing were positive, the UCSF mNGS test yielded the first reported result in 16 (22%) cases. Since most direct detection tests have a lower turnaround time, this result strongly implies that these results were first discovered by UCSF mNGS test and then confirmed by a targeted direct detection test **(Supplementary table 2)**. Among 31 samples with negative mNGS results compared to positive direct detection tests, 14 (45%) were associated with high host background reducing assay sensitivity^10^, 6 (19%) were from patients who had received effective antimicrobial treatment prior to CSF collection, 7 (23%) was from a low titer sample under or near the limit of detection, and 1 (3%) was due to mNGS contamination **(Figure 4)**. Notably, in cases of prior effective antimicrobial treatment, all patients were antigen-positive for either *Cryptococcus sp.* or *Coccidioides sp.* following initiation of treatment for cryptococcal or coccidioidal meningitis, respectively. No clear explanation for discrepant results was found in 3 (10%) false-negative cases.

## Discussion

In this study, we report the analytic results from 7 years of the UCSF mNGS test since the test’s inception in 2016. We found that CSF mNGS testing was the most sensitive test for detection of pathogens in a cohort of patients with clinically severe and diagnostically challenging CNS infections, with 20% of infectious diagnoses only made by the UCSF mNGS test. The UCSF mNGS test was able to identify a broad array of pathogens that are difficult to detect using conventional methods, including novel, emerging, and/or unexpected microorganisms that are not commonly tested for. The positivity rate overall was 14.4% and was higher for UCSF than non-UCSF patients (16.2% vs. 13.9%, p=0.05). This difference seems to be explained by the subthreshold results for UCSF patients that were classified as positives (2.8% vs. 1.8%, p=0.03), where additional clinical information from chart review or communication with the primary physician in challenging cases resulted in reexamination of mNGS results that were consistent with clinical presentation. In the UCSF cohort, most positive subthreshold results (86%) were considered true positive, emphasizing the importance of carefully analyzing results to maximize the UCSF mNGS test yields and reduce the risk of false negative results, especially for high-priority pathogens such as *Mycobacterium tuberculosis* and *Coccidioides immitis*. The analysis of the clinical use of the UCSF mNGS test over 7 years demonstrate its utility and robustness, particularly in cases where routine microbiological testing fails or is less effective: (i) non-culturable, or difficult-to-culture microbes, (ii) vector-borne viruses not commonly tested for, (iii) rare or novel pathogens, (iv) outbreak investigations and (v) “rule-out” of infection.

i. Non-culturable, or difficult-to-culture microbes: The UCSF mNGS test detected many organisms such as fastidious bacterial species that require special incubation conditions for culture or that routinely fail to grow in culture (e.g., *Bartonella henselae*, *Tropheryma whipplei, Borrelia burgdorferi,* and *Leptospira borgpetersenii*). Additionally, the test also detected mycobacteria and fungi that can take weeks to grow in culture, including *Histoplasma capsulatum*, *Aspergillus spp*., *Rhizopus spp*., and *Coccidioides spp.* Importantly, the UCSF mNGS test enabled not only the detection of, but also subtyping of emerging fungal pathogens to the species level, including *Cryptococcus gattii* and *Fusarium solani* (anamorph of *Nectria haematococca*). Lastly, fungal CNS infections, such as from *Histoplasma capsulatum*, are usually difficult to diagnose, requiring a large volume of CSF for culture and/or serologic detection assays performed on multiple sites for successful detection^14^.
ii. Vector-borne viruses: The UCSF mNGS test detected a broad array of vector-borne viruses from the *Flaviviridae* and *Peribunyaviridae* families, such as West Nile virus, Powassan virus, St-Louis virus, and Cache Valley virus. Notably, serology is often considered the gold standard for these types of infections; however, serological results can be challenging to interpret and have limited utility because of their low specificity, long turnaround times, and limited availability in many microbiological laboratories^15,16^. Also, for common pathogens like enteroviruses, the UCSF mNGS test was able to discriminate between species and serotypes in all cases of detection, unlike the typical testing method of NAAT; examples include the detection of enterovirus A71 and enterovirus D68 in cases of meningoencephalitis and/or acute flaccid myelitis^17,18^.
iii. Rare or novel pathogens: The UCSF mNGS test was also useful in the identification of unexpected and/or unusual pathogens in patients with unexplained meningoencephalitis. In many mNGS-positive cases, the clinical presentation and history were atypical, the pathogen was not considered by the treating clinicians *a priori* for testing. Examples include life-threatening amebic infections from *Balamuthia mandrillaris, Acanthamoeba castellanii,* and *Naegleria fowleri*. The availability of diagnostic tests is limited for these parasites, and detection by microscopy or NAAT on CSF is often negative, necessitating a brain biopsy to establish the diagnosis^19^. The UCSF mNGS test could thus alleviate the need for invasive testing. Additional examples of unexpected pathogens include cases of *Candida albicans*, *Toxoplasma gondii*, *Taenia solium* (neurocysticercosis), *Yersinia pestis*, Lymphocytic Choriomeningitis Virus, and *Trypanosoma cruzi* - again, targeted diagnostic tests for these microorganisms are available in specialty referral labs, but were not ordered because there was no *a priori* suspicion.
iv. Outbreak investigations: Results from the UCSF mNGS testing have also been used to initiate outbreak investigations, especially when detecting novel pathogens that have not been described before in human infections. For example, the Potosi virus, a novel *Orthobunyavirus* was detected in mosquitoes from Connecticut^11^, and a vaccine strain of yellow fever virus in CSF from a transplant recipient with encephalitis was recently reported - this case prompted an investigation by the US Centers for Disease Control of a nationwide transplant-associated outbreak^12^. Also, the UCSF mNGS test uncovered an outbreak of fungal meningitis with *Fusarium solani* in the US associated with surgical procedures performed under epidural anesthesia in Matamoros, Mexico^13^.
v. “Rule-out” of infection: In addition to detecting pathogens, the UCSF mNGS test has utility, even with negative results, when infection can be confidently excluded. For example, in complex, undiagnosed cases of meningoencephalitis, the differential diagnosis often includes not only infection, but also autoimmune diseases such as autoantibody syndromes that are treated, often empirically, with steroids and other immunosuppressive agents. Confidently ruling out infection would enable rapid deployment of immunosuppressants and ensure that a yet-to-be-determined infectious etiology does not further progress. In this study, the negative predictive value of CSF mNGS testing was 90.3% (vs 87% for all direct detection tests combined), which is inadequate as a single test to exclude infection. However, the majority (64%) of false negative mNGS results for direct detection from CSF were due either to high DNA host background (45%), a known limitation of mNGS approaches^7,10^, or persistent antigen positivity from fungal infection after onset of treatment (19%). Regarding the latter, it is known that antigens can persist after sterilization in cases of cryptococcal meningitis and that persistent antigen positivity does not correlate with worse clinical outcomes^20^. Thus, in most CSF samples, which do not have high background, a combination of mNGS and other tests such as fungal antigen testing may be sufficient for direct detection of pathogens causing CNS infections in CSF.

This study is different than the prospective PDAID study carried out in 2016 to evaluate the performance and clinical impact of the UCSF mNGS test^6^. In the current study, the UCSF mNGS test is performed as a reference laboratory test with samples sent out from multiple health institutions, and results are reported with minimal clinical information on patients. The inclusion criteria for performing the UCSF mNGS tests are also less stringent (see methods) than in a prospective study with carefully selected populations. Since the publication of the PDAID study, the likelihood of reporting positive subthreshold results has increased, after noting in the study that thresholds were too conservative for some high-priority pathogens present in low abundance in samples. Even with these differences, the sensitivity of the assay is similar between the two studies. When only considering diagnosis made by direct detection of pathogen DNA/RNA on CSF, PPA of mNGS was 80.7% in this study and 80.0% in the PDAID study.

Finally, our study also highlights the limits of the UCSF mNGS test. While the test showed clear utility when compared to direct detection methods on CSF, there are clinical situations where the causative pathogen is absent in CSF and can be found only in brain tissue or well-encapsulated brain abscesses^21^. Additionally, in cases of “hit-and-run” exposure, the window of direct detection in CSF may be brief, such as for West Nile virus meningoencephalitis ^22^. In these cases, diagnostic methods based on detecting pathogens outside of the CNS and/or indirect detection, such as serology, are still useful, and host transcriptome analysis through mNGS appears promising to enhance diagnosis^23,24^.

There are several limitations to our study. Although we report the largest cohort to date evaluating the clinical and diagnostic utility of CSF mNGS in, diagnostically- and clinically-challenging cases of CNS infections, the extent of the diagnostic workup, including types of diagnostics used and clinical decision-making varied widely from case to case, making it difficult to evaluate the side-by-side performance of mNGS compared to other tests, in real-time. Bias may have also been introduced by the use of a composite diagnosis generated from all microbiological test results and clinical adjudication, since a single or consensus gold standard for mNGS has not been established. However, this bias was minimized by conducting three independent assessments. Finally, this study did not evaluate some outstanding questions related to the UCSF mNGS test, including (i) the impact on the clinical management and treatment of patients with CNS infections, (ii) patient characteristics that would maximize diagnostic yield such as transplant, immunocompromised, or pediatric populations, and (iii) the timing and cost-effectiveness of incorporating mNGS into testing algorithms. In today’s clinical workflow, mNGS is often described as the “test of last resort”; however, using mNGS earlier in patient’s clinical course may lead to earlier diagnoses that enable deployment of appropriate and tailored therapy. Ultimately, this might lower healthcare costs, particularly in the in-patient setting where average costs of stays for meningitis/encephalitis cases are over $15,000^25^. The results presented illustrate the clinical utility and yield of including mNGS to the arsenal of diagnostic tools used to investigate and complete the diagnostic workup of patients with suspected CNS infections.

## Methods

### CSF mNGS testing

The UCSF mNGS test was developed to facilitate the broad identification of pathogens in diagnostically challenging CNS infections. The previously reported assay workflow^9^ consisted of (i) nucleic acid extraction, (ii) microbial enrichment using antibody-based removal of methylated host DNA for DNA libraries and DNase treatment for RNA libraries, (iii) library preparation and pooling in equimolar concentrations, and (iv) sequencing on an Illumina (San Diego, California) instrument targeting 5 – 20 million sequences per library. Raw sequences were analyzed using SURPI+, a bioinformatic analysis pipeline for pathogen identification that was modified for clinical use^26^. Automatically generated results from SURPI+ included heat maps of raw and normalized read counts per million (RPM) for each sample and an mNGS results summary in an Excel spreadsheet. A laboratory director (CC or SM) interpreted and reviewed the results before reporting the findings.

Quality control (QC) metrics for the assay included a minimum of 5 million preprocessed reads per sample, >75% of data with quality score >30 (Q>30), and successful detection of all seven representative organisms in the positive control and the internal spiked T1 and MS2 phage controls. A threshold criterion of ≥3 non-overlapping viral reads aligning to the target viral genome was considered a positive detection for virus identification. An RPM ratio threshold of 10 was considered positive for bacteria, fungi, and parasite detection^9^. Following the completion of a prospective study to evaluate clinical utility and diagnostic yield^6^, the reporting algorithm was modified to include additional comments regarding subthreshold detections of likely clinically relevant organisms (for example, *Mycobacterium tuberculosis* and *Coccidioides immitis*) at the discretion of the laboratory director. If the T1 (DNA library) and/or MS2 (RNA library) spiked internal control RPM is <100, indicating high host background, or the number of preprocessed reads is below the established cutoff of 5 million, negative CSF mNGS results are reported with an added comment regarding the potential for decreased sensitivity for organisms with a genome of that library type.

### Patient Cohorts

The UCSF mNGS test results analyzed in this study (n=4,828) included those from samples received from June 2016 to April 2023; samples came from both within and outside the UCSF Health network. Patient demographic information that was available included age, sex, and geographic location by state. Clinical information was available for a subset of cases – those for which the detection of an unusual microorganism prompted a clinical microbial sequencing board (CMSB) discussion with the treating provider and those which were from UCSF patients (“UCSF cohort”). QC metrics and sequencing results were evaluated, along with overall turnaround time (sample collection to result) and laboratory turnaround time (start of processing to result).

The UCSF cohort (n=1,164) from June 2016 to June 2023 was assessed by retrospective chart review to determine the performance and utility in diagnosing CNS infections compared to other assays. All requests for the UCSF mNGS test were reviewed by the laboratory director, and testing was approved if patients met one of more of the following criteria: (i) CSF pleocytosis, (ii) brain biopsy that suggested a likely infectious etiology, (iii) immunocompromised patient with strong suspicion for infection based on clinical presentation, laboratory testing, and/or radiographic imaging, and (iv) recommendation for mNGS testing after neurology and/or infectious diseases consultation. A final composite diagnosis for the UCSF cohort was established through clinical adjudication and review of all diagnostic tests (mNGS and other conventional microbiological testing); this composite diagnosis was performed independently by three infectious disease physicians (PB, MLT and CYC) and categorized results as bacterial, viral, fungal, parasitic, non-infectious/autoimmune, prion-associated, or unknown. Any discrepancies in the final diagnosis were resolved by mutual agreement after re-review and discussion. A case was considered infectious when at least one test result (mNGS or conventional) was positive. All tests relevant to the final diagnosis were recorded for each patient and assigned to 1 of 4 different categories: (1) mNGS; (2) direct detection testing on CSF (culture, antigen detection, and NAAT); (3) direct detection testing sample types other than CSF, such as brain biopsy tissue and/or plasma (culture, antigen detection, and NAAT); and (4) indirect serologic testing from CSF or blood. For each positive test, the turnaround time from sample collection to test result was also recorded. mNGS results were further classified as (i) positive, (ii) single organism with possible contamination, (iii) multiple bacterial/fungal taxa (probable contamination), (iv) incidental findings (organisms detected that are not linked to the CNS infection pathogenesis were not considered a false positive, e.g. detection of HIV detection in an HIV-infected individual), or (v) unclear significance (organisms that were detected and were possibly associated with the infection without any possible confirmation). For the performance analysis, possible and probable contaminants were considered as negative, unless clearly related to the final adjudicated diagnosis. Incidental and unclear significance results were excluded from analysis. A complete review of the adjudication process is available as a **supplementary file**.

## Statistical analysis

Statistical analyses were performed using R Statistical Software (version 4.3.1). Mann-Withney test was used to assess differences in turnaround time. Chi-square testing was used to compare sequencing results yields incorporating individual organism categories. The McNemar test was used to compare sensitivity and specificity differences between diagnostic methods and mNGS testing. Proportional Venn diagrams were constructed using DeepVenn^27^.

## Ethics approval

Retrospective patient chart review and analysis of patient clinical CSF results were performed under a biobanking protocol with waiver of consent approved by the UCSF Institutional Review Board.

## Resource availability

### Lead Contact

Further information and requests for resources and reagents should be directed to and will be fulfilled by the Lead Contact, Charles Chiu.

### Materials Availability

This study did not generate any new reagents.

### Data and Code Availability

The data that support the findings of this study are available on request from the corresponding author C.Y.C. The sequencing data are not publicly available because they contain information that could compromise research participants’ privacy/consent.

## Data Availability

The data that support the findings of this study are available on request from the corresponding author C.Y.C.

## Acknowledgement

We thank the staff at the UCSF Clinical Microbiology Laboratory for performing the UCSF mNGS test. This work was funded, in part, by Delve Bio.

## Authors contribution

P.B., N.B., E.K., V.S., A.P.W., B.O., B.M., S.M and C.Y.C conceived and designed the study. P.B., N.B., M.O., J.N., J.T., M.H., M.L.T., D.I. collected the data. P.B., E.K., V.S., M.L.T., A.P.W., B.O., S.M., and C.Y.C. analyzed data. P.B., E.K., and C.Y.C. wrote the manuscript. P.B., E.K., and C.Y.C. prepared the figures. P.B., N.B., E.K., V.S., M.O., J.N., J.T., M.L.T., C.O., K.Z., M.W., T.B., M.H., D.I., A.P.W., B.O., B.M., S.M., and C.Y.C. edited the manuscript. All authors read the manuscript and agree to its contents.

