## Supplementary table 1 for "Metagenomic next-generation sequencing of cerebrospinal fluid for diagnosis of central nervous system infections: 7-year performance of a clinically validated test"

| Non-fastidious pathogens | N | Uncommon and/or difficult to diagnose pathogens | N |
| --- | --- | --- | --- |
| *Streptococcus spp.* | 24 | *Mycobacterium tuberculosis* | 13 |
| *Klebsiella spp.* | 12 | *Mycobacterium non-tuberculosis* | 7 |
| *Staphylococcus spp.* | 9 | *Nocardia farcinia* | 3 |
| *Enterococcus spp.* | 8 | *Borrelia burgdorferi* | 2 |
| *Haemophilus spp.* | 7 | *Mycoplasma hominis* | 2 |
| *Neisseria spp.* | 4 | *Treponema pallidum* | 2 |
| *Serratia spp.* | 4 | *Ureaplasma parvum* | 2 |
| *Citrobacter spp.* | 3 | *Actinomyces oris* | 1 |
| *Cutibacterium spp.* | 2 | *Bartonella henselae* | 1 |
| *Enterobacter spp.* | 2 | *Borrelia miyamotoi* | 1 |
| *Pseudomonas spp.* | 2 | *Chlamydia pstittaci* | 1 |
| *Acinetobacter spp.* | 1 | *Cronobacter sakazakii* | 1 |
| *Bacteroides spp.* | 1 | *Fusobacterium necrophorum* | 1 |
| *Corynebacterium spp.* | 1 | *Fusobacterium nucleatum* | 1 |
| *Cronobacter spp.* | 1 | *Gardenerella vaginalis* | 1 |
| *Leuconostoc spp.* | 1 | *Kingella kingae* | 1 |
| *Moraxella spp.* | 1 | *Legionella sp.* | 1 |
| *Pantoea spp.* | 1 | *Leptospira borgpetersenii* | 1 |
| *Proteus app.* | 1 | *Mycoplasma pneumoniae* | 1 |
| *Rothia spp.* | 1 | *Tropheryma whipplei* | 1 |
| *Shewanella spp.* | 1 | *Ureaplasma urealyticum* | 1 |
|  |  | *Yersinia pestis* | 1 |

**Supplementary table 1. Bacteria detection list by mNGS**
