## Supplementary table 2 for "Metagenomic next-generation sequencing of cerebrospinal fluid for diagnosis of central nervous system infections: 7-year performance of a clinically validated test"

**Supplementary table 2. Diagnosis made by mNGS**

| **Diagnosis made by mNGS but negative by direct detection methods (n=58)** | | | | |
| --- | --- | --- | --- | --- |
| **DNA virus** | **RNA virus** | **Bacteria** | **Fungi** | **Parasite** |
| Cytomegalovirus | Colorado tick fever virus | *Enterobacter cloacae* | *Aspergillus sp.* (5) | Acanthamoeba castellani |
| Epstein-Barr virus (2) | Coxsackievirus A6 | *Fusobacterium necrophorum* | *Coccidioides immitis* (2) | *Balamuthia mandrillaris* (2) |
| HHV-7 (2) | Coxsackievirus B5 (3) | *Klebsiella aerogenes* | *Mucorales sp.* | *Toxoplasma gondii* (5) |
| HHV-6 (2) | Echovirus 30 | *Mycobacterium bovis* |  |  |
| JC virus | Echovirus 6 | *Mycobacterium tuberculosis* (2) |  |  |
| Varicella Zoster Virus (2) | LCMV | *Neisseria meningitidis* |  |  |
| WU Polyomavirus | West Nile Virus (5) | *Nocardia nova* |  |  |
|  |  | Polymicrobial |  |  |
|  |  | *Streptococcus agalactiae* |  |  |
|  |  | *Streptococcus intermedius* |  |  |
|  |  | *Streptococcus pneumoniae* (2) |  |  |
|  |  | *Treponema pallidum* (2) |  |  |
|  |  | *Tropheryma whipplei* |  |  |
|  |  | *Ureaplasma parvum* (2) |  |  |
| **Diagnosis made faster by mNGS than direct detection methods when both were positive (n= 16/72, 22%)** | | | | |
| Cytomegalovirus (2) | Enterovirus A71 | *Mycobacterium tuberculosis* | *Aspergillus sp.* (2) | *Acanthamoeba sp.* |
| Epstein-Barr virus | West Nile Virus | *Nocardia farcinia* | *Coccidioides immitis* (2) | *Angiostrongylus cantonensis* (2) |
|  |  | *Streptococcus pneumoniae* | *Fusarium sp.* |  |
