## Supplementary table 3 for "Metagenomic next-generation sequencing of cerebrospinal fluid for diagnosis of central nervous system infections: 7-year performance of a clinically validated test"

| **Final causative organism** | **Testing modality positive** | **Infection category** | **Discrepant factor when mNGS negative and Direct CSF detection positive** |
| --- | --- | --- | --- |
| *Acanthamoeba sp.* | PCR | Parasitic | High human background |
| *Angiostrongylus cantonensis* | PCR | Parasitic | High human background |
| *Aspergillus fumigatus* | uPCR | Fungal | High human background |
| *Bordetella hinzii* | Cx | Bacterial | High human background |
| *Candida parapsilosis* | Cx | Fungal | High human background |
| CMV | PCR | DNA virus | low positive sample |
| *Coccidioides sp.* | Ag | Fungal | Treated before CSF collection |
| *Coccidioides sp.* | Cx | Fungal | High human background |
| *Coccidioides sp.* | Ag | Fungal | Treatment before CSF testing |
| *Cryptococcus neoformans* | Ag | Fungal | low positive sample |
| *Cryptococcus neoformans* | Ag | Fungal | Treated before CSF collection |
| *Cryptococcus neoformans* | Ag | Fungal | Treated before CSF collection |
| *Cryptococcus neoformans* | Ag | Fungal | High human background |
| *Cryptococcus neoformans* | Cx | Fungal | low positive sample (CrAg-) |
| *Cryptococcus neoformans* | Ag | Fungal | Treated before CSF collection |
| *Cryptococcus neoformans* | Ag/Cx | Fungal | Treated before CSF collection |
| *Cryptococcus neoformans* | Ag/Cx | Fungal | High human background |
| *Cutibacterium acnes* | Cx | Bacterial | High human background |
| *Cutibacterium acnes* | Cx | Bacterial | Unknown |
| EBV | PCR | DNA virus | low positive sample |
| *Escherichia coli* | Cx | bacterial | High human background |
| HSV | PCR | DNA virus | High human background |
| HSV-2 | PCR | DNA virus | High human background |
| *Mycobacterium tuberculosis* | Cx | Bacterial | High human background |
| Parvovirus B19 | PCR | DNA virus | low positive sample |
| *Scedosporium apiospermum* | Cx | Fungal | low positive sample (CrAg-) |
| *Staphylococcus aureus* | Cx | Bacterial | High human background |
| *Staphylococcus capitis* | Cx | bacterial | Unknown |
| *Steprococcus agalactiae* | uPCR | Bacterial | mNGS Contamination with other bacteria |
| *Taenia sp.* | Ag | Parasitic | Unknown |
| VZV | PCR | DNA virus | low positive sample |

**Supplementary table 3. False negative mNGS results**
