## Supplementary table 4 for "Metagenomic next-generation sequencing of cerebrospinal fluid for diagnosis of central nervous system infections: 7-year performance of a clinically validated test"

**Supplementary table 4. False positive mNGS results**

| **mNGS false positive results description (n=11)** | | | | |
| --- | --- | --- | --- | --- |
| **mNGS result** | **Diagnosis category** | **Action upon result** | **Compatible with clinical presentation** | **Clinical description** |
| *Corynebacterium sp.* | Unknown | No | No | Meningoencephalitis with response to antituberculous therapy. |
| *Shewanella baltica* | Unknown | No | No | CNS enhancing lesions of unknown etiology that resolved without antibacterial treatment. |
| *Leuconostoc sp.* | AINI | No | No | Diagnosis of autoimmune demyelinating disease, with brain biopsy not compatible with an infectious process. |
| Varicella Zoster virus | Unknown | No | No | Encephalopathy with mild CSF pleiocytosis that resolved without antiviral treatment. |
| *Pantoea sp.* | Unknown | No | No | Facial myokymia with no pleiocytosis in CSF. |
| *Staphyloccoccus epidermidis* | Unknown | No | No | Status epilepticus of unknown etiology with no pleiocytosis in CSF. Target detected not compatible with clinical presentation. |
| *Cutibacterium avidum* | Unknown | No | No | Patient already undergoing treatment for possible fungal meningoencephalitis with subsequent improvement. Target detected not compatible with clinical presentation. |
| *Mycoplasma hominis* | Unknown | No | No | Nosocomial meningitis with improvement without effective antibiotics against Mycoplasma hominis. |
| Dengue virus, *Candida parapsilosis* | AINI | No | No | Diagnosis of CNS neoplasic metastasis. Target detected not compatible with clinical presentation and epidemiology. |
| *Staphylococcus aureus* | AINI | No | No | Diagnosis of diffuse idiopathic arachnoiditis. Target detected not compatible with clinical presentation. |
| *Enterococcus faecalis* | Unknown | No | No | Diagnosis of idiopathic myelitis. Target detected not compatible with clinical presentation. |
