## Supplementary table 5 for "Metagenomic next-generation sequencing of cerebrospinal fluid for diagnosis of central nervous system infections: 7-year performance of a clinically validated test"

Supplementary table 5: Detection of single organism with possible contamination on report.

**Detection of a target with a comment for possible contamination on mNGS that are true positive (n=2/22, 9.1%)**

|  | **mNGS result** | **Diagnosis category** | Bacterial or fungal pathogen compatible with clinical presentation | **Clinical description** |
| --- | --- | --- | --- | --- |
| 1 | *Corynebacterium sp.* | Bacterial | Yes | Meningitis in a patient with a ventriculo-peritoneal shunt. CSF culture also positive for Corynebacterium sp. |
| 2 | *Staphylococcus aureus* | Bacterial | Yes | Nosocomial meningitis in a patient post neurosurgery. CSF culture also positive for Staphylococcus aureus. |
| 3 | *Pantoea spp.* | Auto-immune/Non-infectious | No |  |
| 4 | *Staphylococcus haemolyticus* | Auto-immune/Non-infectious | No |  |
| 5 | *Stenotrophomonas rhizophila* | Bacterial (*Borrelia burgdorferi*) | No |  |
| 6 | *Deinococcus proteolyticus* | Auto-immune/Non-infectious | No |  |
| 7 | *Lactobacillus reuteri* | Auto-immune/Non-infectious | No |  |
| 8 | ***Balamuthia mandrillaris****, Micrococcus sp.* | Parasitic (*Balamuthia mandrillaris*) | No |  |
| 9 | *Candida tropicalis* | Auto-immune/Non-infectious | No |  |
| 10 | *Pseudomonas sp.* | Auto-immune/Non-infectious | No |  |
| 11 | *Propionibacterium sp* | unknown | No |  |
| 12 | *Cutibacterium avidum* | unknown | No |  |
| 13 | *Corynebacterium spp* | Auto-immune/Non-infectious | No |  |
| 14 | *Bacillus infantis* | unknown | No |  |
| 15 | *Sphingomonas sp.* | unknown | No |  |
| 16 | *Staphylococcus warneri* | unknown | No |  |
| 17 | *Clostridium botulinum* | unknown | No |  |
| 18 | *Lactobacillus delbrueckii* | unknown | No |  |
| 19 | *Pantoaea ananatis* | DNA virus (VZV) | No |  |
| 20 | *Enterobacter cloacae* | unknown | No |  |
| 21 | *Staphylococcus warneri* | Auto-immune/Non-infectious | No |  |
| 22 | *Staphylococcus hominis* | Auto-immune/Non-infectious | No |  |
