## Supplementary table 6 for "Metagenomic next-generation sequencing of cerebrospinal fluid for diagnosis of central nervous system infections: 7-year performance of a clinically validated test"

Supplementary table 6: Detection of multiple bacterial/fungal taxa

**Detection of multiple bacterial/fungal taxa on mNGS that are true positive (n=1/65, 1.5%)**

|  | **mNGS result** | **Diagnosis category** | **Multiple taxa detection compatible with clinical presentation** | **Clinical description** |
| --- | --- | --- | --- | --- |
| 1 | *Porphyromonas gingivalis, Prevotella dentalis, Streptococcus milleri* | Bacterial | Yes | Odontogenic infection with bilateral sinus cavernous thrombosis with neutrophilic pleocytosis and negative CSF culture. |
| 2 | *Micrococcus luteus, Pantoea sp.* | unknown | No |  |
| 3 | *Rothia dentocariosa, Streptococcus sanguinis,and Streptococcus parasanguinis* | Auto-immune/Non-infectious | No |  |
| 4 | *Morganella morganii* and others | unknown | No |  |
| 5 | *Sphingomonas sp, Clavibacter sp* | unknown | No |  |
| 6 | *Bifidobacterium dentiuym, Streptococcus mutans, Methylobacterium populi* | Auto-immune/Non-infectious | No |  |
| 7 | *Pantoea spp., Corynebacterium ureicelerivorans* | Bacterial (*Streptococcus agalactiae*) | No |  |
| 8 | *Prevotella, Porphyoromonas, Streptococcus, Haemophilus spp* | Auto-immune/Non-infectious | No |  |
| 9 | *Pantoea spp, Bifidobacterium thermophilum* | Auto-immune/Non-infectious | No |  |
| 10 | *Pseudomonas putida* and others | unknown | No |  |
| 11 | *Bacillus megaterium, Rhodococcus equi* | unknown | No |  |
| 12 | *Corynebacterium spp* and others | unknown | No |  |
| 13 | Multiple bacterial taxa | unknown | No |  |
| 14 | *Klebsiella pneumoniae, Enterobacter cloacae, Citrobacter freundii* | unknown | No |  |
| 15 | *Lactobacillus acidophilus* and others | Auto-immune/Non-infectious | No |  |
| 16 | *Streptococcus agalactiae, Exiguobacterium sp., HIV.1,* ***Cryptococcus neoformans*** | Fungal (C. neoformans) | No |  |
| 17 | *Prevotella melanogenica, Streptococcus sp* | Auto-immune/Non-infectious | No |  |
| 18 | Multiple bacterial taxa | unknown | No |  |
| 19 | *Pseudomonas simiae ,Streptococcus salivarius, Bacteroides vulgatus* | Auto-immune/Non-infectious | No |  |
| 20 | *Geobacillus sp.,Enterobacter sp.,and Leuconostoc sp., Candida famata* | unknown | No |  |
| 21 | *Corynebacterium urealyticum* and others | unknown | No |  |
| 22 | *Lactobacillus, Pantoea, Moraxella catarrhalis* | unknown | No |  |
| 23 | *Enterobacter cloacae, Rothia dentocariosa,* **HIV** | RNA virus (HIV) | No |  |
| 24 | *Chroococcidiopsis thermalis* | unknown | No |  |
| 25 | *Streptococcus lutitiensis, Deinococcus proteolyticus, Lactobacillus animalis* | Auto-immune/Non-infectious | No |  |
| 26 | *Cutibacterium acnes* and others | Auto-immune/Non-infectious | No |  |
| 27 | Multiple bacterial taxa | unknown | No |  |
| 28 | *Blastococcus saxobsidens, Geodermatophilus obscurus, Bifidobacterium thermophilum, Granulibacter bethesdensis, Modestobacter marinus,* **CMV** | DNA virus (CMV) | No |  |
| 29 | *Micrococcus luteus, Streptococcus sp., Dietzia sp* | Auto-immune/Non-infectious | No |  |
| 30 | *Megamonas hypermegale, Lactobacillus reuteri, Anoxybacillus flavithermus* | Auto-immune/Non-infectious | No |  |
| 31 | *Modestobacter marinus, Azospirilllum lipoferum, Spirosoma linguale, Variovarox paradoxus* | Auto-immune/Non-infectious | No |  |
| 32 | *Staphylococcus aureus, Staphylococcus epidermidis, Micrococcus luteus, Serratia marcescens* | unknown | No |  |
| 33 | *Penicillium rubrens* and others | Auto-immune/Non-infectious | No |  |
| 34 | *Bacillus licheniformis, Micrococcus luteus, Gordonia bronchialis* | unknown | No |  |
| 35 | *Streptococcus thermophilus, Bifidobacterium animalis, Corynebacterium ureicelerivorans* | Auto-immune/Non-infectious | No |  |
| 36 | *Micrococcus luteus, Klebsiella pneumoniae, Kytococcus sendentarius* | Auto-immune/Non-infectious | No |  |
| 37 | *Trichophyton rubrum* and others | unknown | No |  |
| 38 | *Lactobacillus sakei, Klebsiella pneumoniae, Acidovorax sp., Rhodococcus erythropolis* | Auto-immune/Non-infectious | No |  |
| 39 | *Gardnerella vaginalis, Corynebacterium sp., Staphylococcus auricularis* | Auto-immune/Non-infectious | No |  |
| 40 | *Staphylococcus pettenkoferi, Mycobacterium paragordonae, Bacillus coagulans, Neisseria sicca, Rothia mucilaginosa* | unknown | No |  |
| 41 | *Mycobacterium choelonae* and others | unknown | No |  |
| 42 | *Bifidobacterium adolescentis* and others | unknown | No |  |
| 43 | *Micrococcus, Pseudomonas, Corynebacterium sp,* ***Cryptococcus neoformans,*** JC polyomavirus, HIV1 | Fungal (C. neoformans) | No |  |
| 44 | *Acinetobacter radioresistens, Sphingomonas sp., Rhizobium sp.,Stenotrophomonas sp.* | Auto-immune/Non-infectious | No |  |
| 45 | *Corynebacterium sp., Staphylococcus sp., Malassezia sp., Anthracocystis sp, Tilletiopsis sp.* | Auto-immune/Non-infectious | No |  |
| 46 | *Porphyomonas sp.,Corynebacterium sp., Stenotrophomonas sp.* | Auto-immune/Non-infectious | No |  |
| 47 | *Acinetobacter lwoffii, Rothia mucilaginosa, Staphylococcus sp.* | unknown | No |  |
| 48 | *Staphylococcus sp., Corynebacterium sp., Rothia mucilaginosa* | unknown | No |  |
| 49 | *Streptococcus thermophilus, Gardnerella vaginalis, Lactobacillus plantarum, Cutibacterium acnes* | Auto-immune/Non-infectious | No |  |
| 50 | *Bacteroides sp., Enterococcus faecium, Corynebacterium sp., Cutibacterium acnes* | Auto-immune/Non-infectious | No |  |
| 51 | *Enterococcus cecorum, Chryseobacterium haifense, Bacillus flexus* | unknown | No |  |
| 52 | *Neisseria sp,Corynebacterium sp., Haemophilus parainfluenzae* | Auto-immune/Non-infectious | No |  |
| 53 | *Pseudomonas fluorescens, Prevotella sp., Bacteroides vulgatus* | unknown | No |  |
| 54 | *Acineotbacter sp.,Capnocytophaga sp., Lactobacillus crispatus* | unknown | No |  |
| 55 | *Staphylococcus pettenkoferi,Chryseobacterium haifense, Corynebacterium sp.* | Fungal (*Coccidioides sp.*) | No |  |
| 56 | *Corynebacterium striatum, Serratia grimesii* | unknown | No |  |
| 57 | *Staphylococcus cohnii,Chryseobacterium haifense, Gordonia bronchialis* | Auto-immune/Non-infectious | No |  |
| 58 | *Pseudomonas sp., Capnocytophaga sp., Bifidobacterium thermophilum* | unknown | No |  |
| 59 | *Lactobacillus crispatus, Propionibacterium sp., Staphylococcus aureus* | Auto-immune/Non-infectious | No |  |
| 60 | *Haemophilus parainfluenzae, Pseudomonas sp., Acinetobacter radioresistens* | unknown | No |  |
| 61 | *Neisseria subflava,   Rothia mucilaginosa, and Streptococcus thermophilus,* *Penicillium rubens, Aspergillus glaucus* | unknown | No |  |
| 62 | Multiple bacterial taxa, HIV1, ***Cryptococcus neoformans*** | Fungal (*C. neoformans*) | No |  |
| 63 | *Bacteroides spp* and others | unknown | No |  |
| 64 | *Enterococcus faecalis, Lactobaillus crispatus, Bifidobacterium dentium, Candida famata* | Auto-immune/Non-infectious | No |  |
| 65 | *Rhodococcus sp., Enterococcus cecorum, Staphylococcus hominis, Pseudomonas koreensis,Sphingomonas taxi* | Auto-immune/Non-infectious | No |  |
